# Conservative surgery vs hysterectomy for placenta accreta spectrum disorders: a systematic review and meta-analysis

**DOI:** 10.1101/2025.02.04.25321691

**Authors:** Gabriel Moreira Lino, Pauliana Valéria Machado Galvão, Valda Lúcia Moreira Luna, George Alessandro Maranhão Conrado

**Affiliations:** Universidade de Pernambuco, Serra Talhada, Brazil; Faculdade de Ciências Médicas, Universidade de Pernambuco, Recife, Brazil

**Author notes:** Corresponding author: **Pauliana Valéria Machado Galvão,** Address: R. Arnóbio Marquês, 310 - Santo Amaro, Recife - PE, 50100-130.

**Keywords:** Placenta accreta, Surgery, Fertility, One-step surgery, Triple-P procedure, Conservative management

## Abstract

**Objective:** Resection and reconstruction of the uterus may be an alternative to hysterectomy in placenta accreta spectrum disorders. We aimed to review whether surgical and maternal outcomes were comparable between these techniques.

**Data sources:** Medline, Embase, and the Cochrane Library databases were searched until 1 January 2025.

**Study selection:** We included randomized controlled trials (RCT) and nonrandomized studies (NRSI) comparing conservative surgery, involving resection and reconstruction of the myometrium without placental removal, to hysterectomy in patients with placenta accreta, increta, or percreta.

**Data collection:** Two authors were responsible for independently screening the title and abstract, and the full texts of all identified records. We assessed the risk of bias with ROB-2 or ROBINS-I, and the quality of evidence with GRADE. The main analysis only included studies without a high risk of bias. A secondary non-inferiority Bayesian analysis was reported alongside the results.

**Data synthesis:** We included one RCT and eleven nonrandomized studies (1586 women). Compared to hysterectomy, conservative surgery may result in a similar number of transfused packs of red blood cells (MD −0.70 packs, 95% CI −1.28 to −0.12; 1 RCT and 4 NRSI, 649 women; I^2^= 0%; low-certainty evidence), and a similar risk of bladder injury (RR 0.36, 95% CI 0.12 to 1.07; 1 RCT and 4 NRSI, 649 women; I^2^= 0%; low-certainty evidence). Excluding rare outcomes, the probability that conservative surgery is non-inferior to hysterectomy was greater than 90.0%.

**Conclusions:** On the basis of low evidence, conservative surgery did not increase the risk of clinically important harm compared to hysterectomy, with the benefit of uterus preservation. Further RCTs are needed to reach definitive conclusions.

**Registration:** PROSPERO CRD42024601117.

## INTRODUCTION

Until recently, hysterectomy was virtually the only management option for placenta accreta spectrum disorder (PAS). However, as the prevalence of PAS has increased to affect approximately 1 in every 313 women undergoing cesarean section, the growing number of women wishing to preserve fertility has prompted investigation into alternative strategies for uterine preservation^1^. Currently, leaving the placenta in situ is one of the most utilized alternatives to hysterectomy, although it requires high-resource settings and continuous long-term monitoring^2,3^. In this context, uterus-preserving surgery (also referred to as one-step surgery and triple P procedure, according to variations in the technique proposed by individual authors) has been developed to provide immediate surgical resolution of PAS during cesarean delivery^4^.

This technique involves removing the invaded myometrium and reconstructing the uterine wall, with or without preventive surgical devascularization^2,4^. However, it remains to be proven whether surgical outcomes are superior or similar to hysterectomy, specifically because of the challenging surgical anatomy and the inherent risk of intrapartum hemorrhage and visceral organ injury. Therefore, this review aims to summarize the available evidence on this conservative surgical technique, comparing it to hysterectomy in patients diagnosed with PAS who wish to preserve fertility.

## METHODS

### Protocol and registration

This systematic review was registered prospectively in the PROSPERO database, identifier CRD42024601117. We followed the PRISMA 2020 guidelines for the reporting of systematic reviews and meta-analyses^5^.

### Criteria for considering studies for this review

We considered randomized controlled trials (RCT) and nonrandomized studies of interventions comparing conservative surgery to hysterectomy in patients with placenta accreta, increta, or percreta. To be included, the surgical technique must have involved resection of abnormally invasive placenta and myometrium and reconstruction of the uterine wall. Therefore, we excluded studies that provided fertility preservation by manual removal of the placenta followed by vascular control procedures, such as uterine compressive sutures, tourniquets, and uterine devascularization.

The following critical outcomes were considered: intraoperative blood loss, number of transfused packs of red blood cells or women needing transfusion, intraoperative visceral organ injury (bladder injury, bowel injury, ureter injury, and vascular injury), reoperation, long-term complications of surgery (fistula, premature ovarian insufficiency, urinary dysfunction, dysmenorrhea). The secondary outcomes of the review were: duration of surgery, duration of hospital stay, admission to the intensive care unit, and postoperative infection. Studies had to report at least one of the primary outcomes to be eligible for the review.

### Search methods for identification of studies

We searched for published studies up to January 2025, on Medline, Embase, and the Cochrane Library using the terms: (one step conservative surgery OR uterine conservative-resective surgery OR uterus resection OR uterine resective surgery OR uterine conservative surgery OR uterus conservation surgery OR uterine preserving surgery OR fertility sparing surgery OR fertility preserving surgery OR triple p procedure OR triple p) and (placenta accreta spectrum OR placenta increta OR placenta percreta OR abnormally invasive placenta OR morbidly adherent placenta). There were no language restrictions.

### Data collection and analysis

Two authors were responsible for independently screening the title and abstract of all identified records. When one of the authors considered the record eligible, both assessed the full text. Thereafter, they proceeded with selecting and extracting information about study characteristics, which were summarized on a cross-validated file.

### Risk of bias assessment in included studies

For the risk of bias assessment, two authors independently used the Cochrane risk-of-bias tool for randomized trials (RoB 2), version of August 2019^6^. The ROBINS-I V2 tool, 2024, was used instead for nonrandomized studies of interventions^7^. We evaluated the overall risk of bias in each study based on three primary outcomes (blood loss, transfusion, and intraoperative visceral organ injury). Dropouts, group conversions, or loss to follow-up below 5% were judged as low risk of bias.

### Synthesis methods

We used an inverse-variance random effects model, reporting dichotomous data as Relative Risk (RR) with 95% Confidence intervals (CI) and continuous outcomes as mean differences (MD) and 95% CI. Statistical heterogeneity was assessed by calculating tau (DerSimonian-Laird estimator and Jackson method for confidence interval of tau^2^), I², and Cochran’s Q; visual I² variation that included clinically important differences indicated serious heterogeneity^8^. We investigated publication bias with funnel plots and Egger’s test.

Studies that reported medians and interquartile ranges were transformed using an internal function of the meta package, following what was described by Luo and collaborators^9^. We stratified our analyses based on the risk of bias. Primary results in every section only included studies with randomized design or that were not at high risk of bias. Subsequent analyses reported the estimated effect including the remaining studies, except when the results of both analyses were uncertain, making it unnecessary to report them separately.

Analyses were conducted in R, with the packages meta and metafor^10,11^.

### Sensitivity analyses

To explore the certainty of the main results, we conducted a planned secondary analysis using a Bayesian framework. Specifically, we assessed how integrating prior knowledge from observational studies not at serious or critical risk of bias would influence the interpretation of the randomized studies, which had limited statistical power due to modest sample sizes.

We first synthesized the selected nonrandomized studies using a Bayesian meta-analysis with a non-informative prior (mean = 0, SD = 10,000), generating a posterior distribution that served as an informative prior for the subsequent analysis of the trial data. We then combined this prior with the likelihood derived from the randomized controlled trial to obtain a posterior estimate reflecting both sources of evidence.

This reanalysis focused on critical clinical outcomes: number of packs of red blood cells transfused, intraoperative visceral injuries (including bladder and ureter), need for reoperation, and long-term surgical complications.

### Certainty of the evidence assessment

The overall certainty of the evidence for each outcome was assessed by two authors using the Grading of Recommendations, Assessment, Development and Evaluation (GRADE) handbook^12^. The quality of the evidence was summarized in a Summary of Findings table generated in GRADEpro GDT. For the evaluation of imprecision, the update provided in 2022 was followed^13^. The Review Information Size was determined using conventional power analysis (α = 0.05; β = 0.2), taking into account the range of standard deviations from randomized controlled trials and medium to low risk of bias studies.

## RESULTS

### Results of the search

We initially identified 640 records in our search. Of these, 106 were deemed potentially eligible, and 12 (only one RCT) were ultimately included in the review. The primary reasons for exclusion were inadequate surgical techniques (manual placental removal or failure to report surgical methods), study design (such as case series, reports, reviews, editorials, and letters), absence of relevant outcome data specified in the review protocol, and not comparing conservative surgery to hysterectomy (such as comparing to manual removal of the placenta).

### Included studies

Table 1 summarizes the characteristics of the included studies. One RCT and eleven non-randomized studies were included, four of which had prospective designs. Moreover, some studies reported the PAS FIGO clinical grading^14^, with patients of every grade in both arms. The average maternal age ranged from 30 to 37 years, with most women having had one or two previous cesarean sections.

**Table 1.**
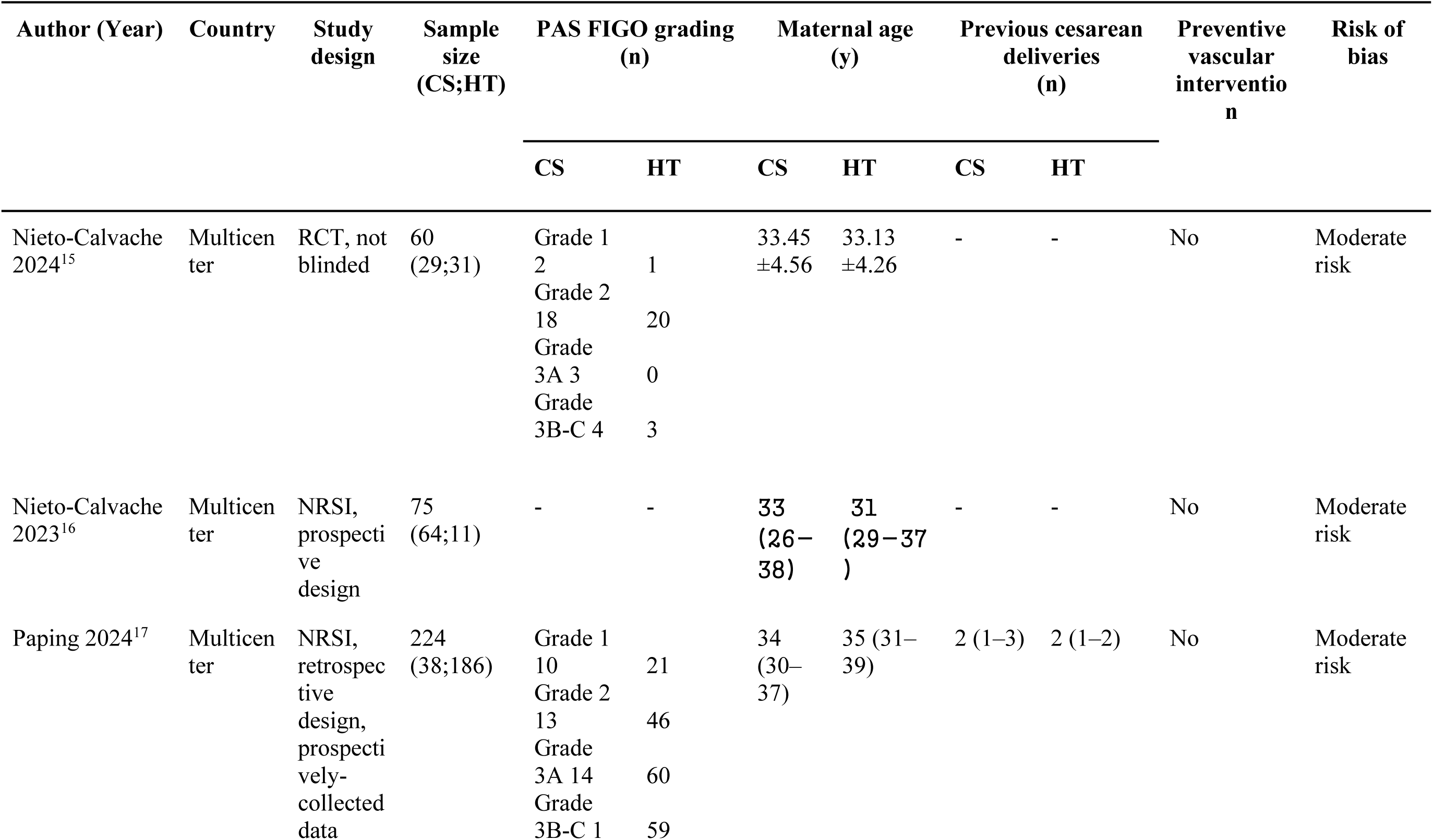
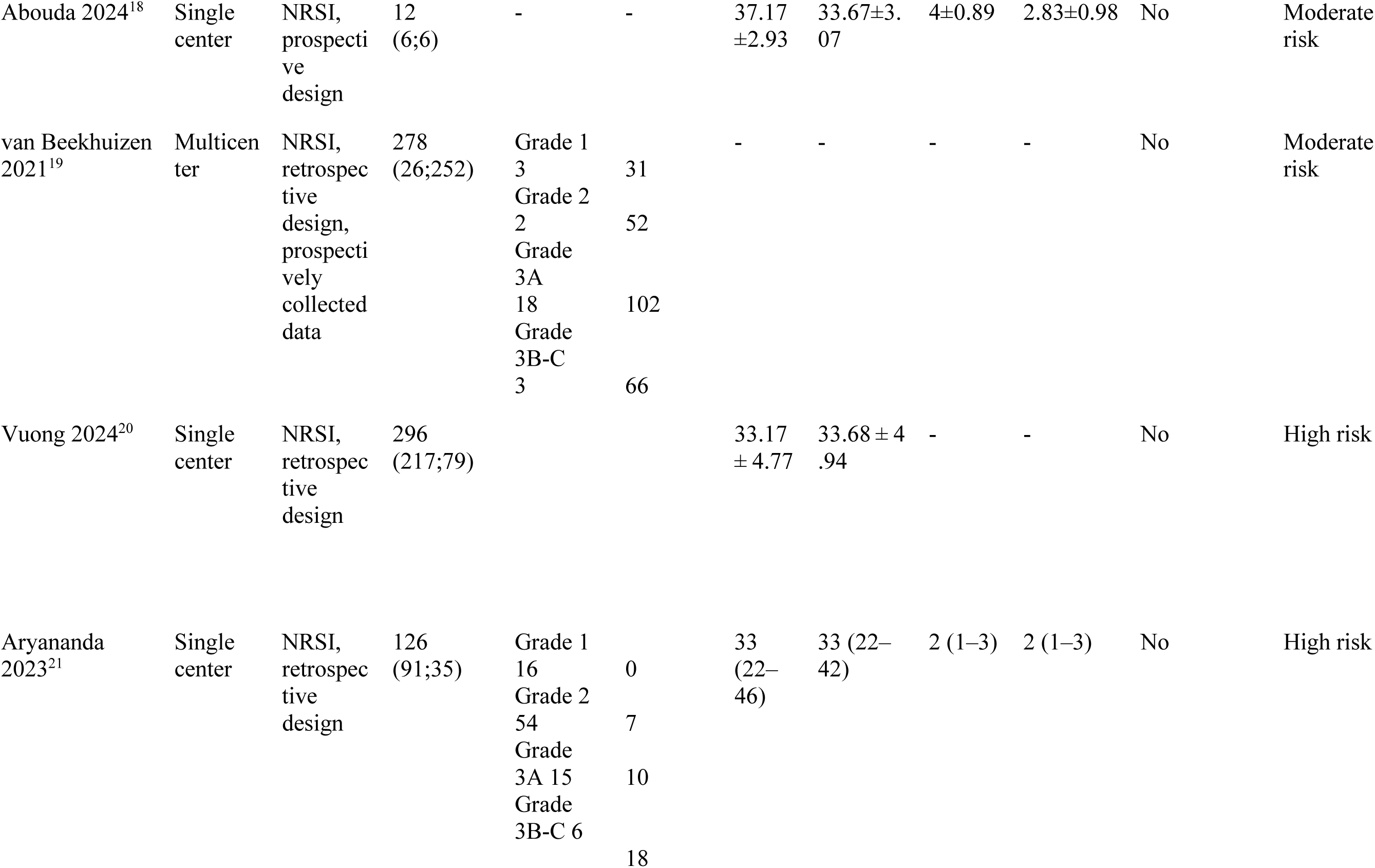
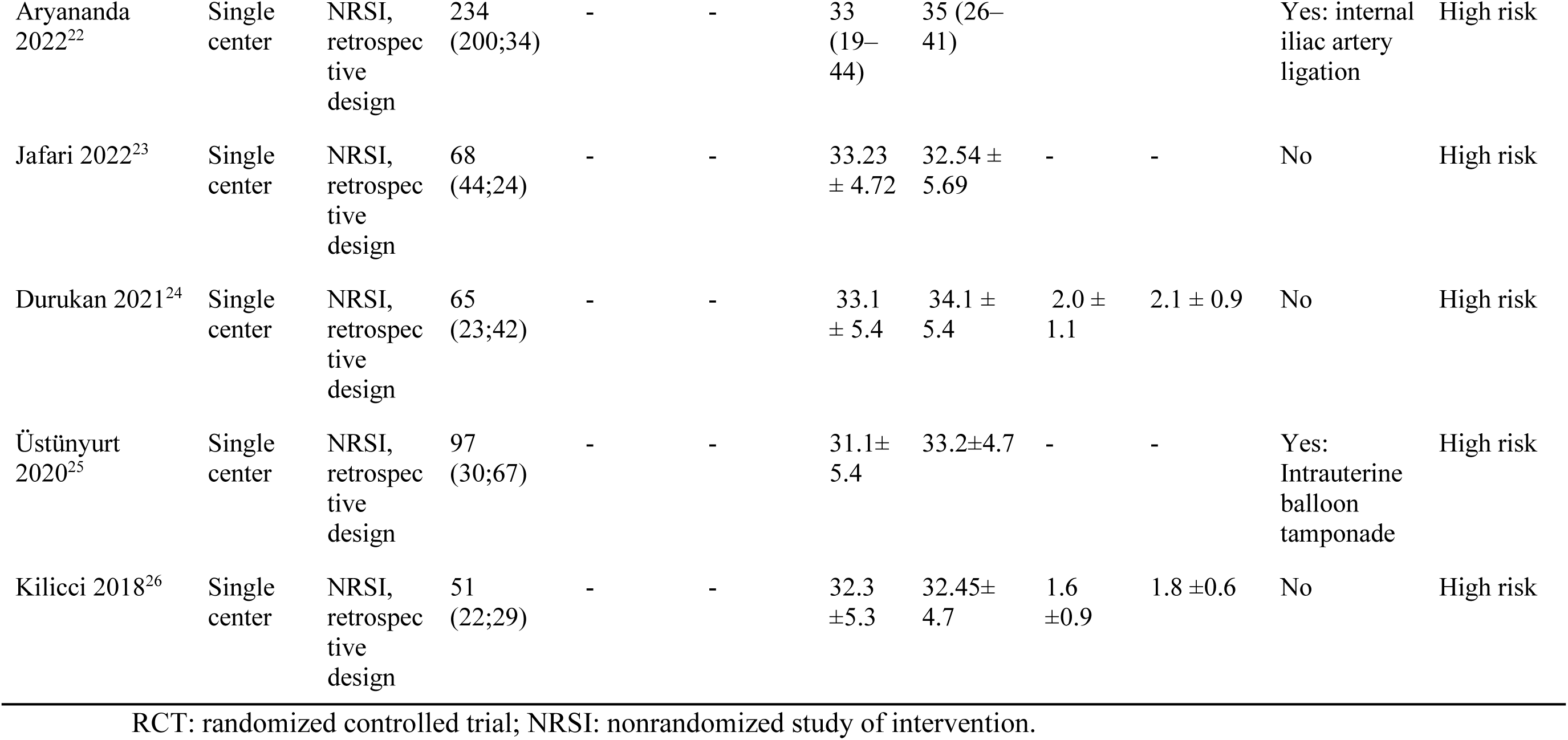
Summary of characteristics of included studies.

Preventive vascular interventions before myometrial resection were utilized in three of the included studies. Further descriptions of methods, eligibility criteria, and participants are available in Supplementary Table S1. In general, the most common eligibility criteria for uterine conservative surgery were: (1) at least 2 cm of healthy myometrium in the anterior uterine wall, (2) more than 50% of the uterine circumference without PAS involvement, and (3) the ability to safely dissect the bladder from the uterus. Most studies followed the surgical technique described as one-step surgery^4^.

### Risk of bias of included studies

An overview of the risk of bias is provided in Table 1, and the detailed description is presented in Table S2. The included RCT was classified as having a moderate risk of bias due to the lack of blinding and important patient crossover/drop-out. Nearly all non-randomized studies were considered at high risk of bias, as patient selection was often at serious risk of bias. Additionally, few studies reported potential confounding variables or adjusted their analyses as necessary.

An investigation into publication bias revealed that retrospective studies, which were at high risk of bias, generally reported higher mean differences with larger standard errors compared to studies with moderate to low risk of bias (Figure S1). However, Egger’s Test did not detect significant publication bias affecting the analysis (p-value = 0.20).

### Synthesis of results

#### 1.1 Intraoperative blood loss

Conservative surgery may result in similar intraoperative blood loss compared to hysterectomy (MD −320.89 mL, 95% CI −574.95 to −66.82; 1 RCT and 3 NRSI, 637 women; I^2^= 14%; low-certainty evidence). By including studies at high risk of bias, the difference in blood loss is larger, but of lower certainty (MD −497.33 mL, 95% CI −755.37 to −226.17; 1 RCT and 7 NRSI; I^2^= 73%; very low-certainty evidence)(Figure 2).

**Figure 1.** PRISMSA 2020 flow diagram of study selection

**Figure 2.** Comparison of uterine conservative surgery vs hysterectomy on intraoperative blood loss in women with placenta accreta spectrum disorders

#### 1.2 Number of transfused packs of red blood cells (pRBCs) and number of women needing transfusion

The number of transfused pRBCs per patient may be similar in conservative surgery compared to hysterectomy (MD −0.70 packs, 95% CI −1.28 to −0.12; 1 RCT and 4 NRSI, 649 women; I^2^= 0%; low-certainty evidence). This benefit could be slightly larger (MD −1.15 packs, 95% CI −1.64 to −0.65; 1 RCT and 9 NRSI; I^2^= 20%; very low-certainty evidence)(Figure 3). Further, assuming a non-inferiority margin of 0.5 pRBCs, the probability that conservative surgery is non-inferior to hysterectomy is 95.4% (Figures S2 and S3).

**Figure 3.** Comparison of uterine conservative surgery to hysterectomy on the number of transfused packs of red blood cells per patient in women with placenta accreta spectrum disorders. RCT, randomized controlled trial; IV, inverse variance; NRSI, nonrandomized study of intervention.

Moreover, there may be a similar number of patients needing blood transfusion (RR 0.87, 95% CI 0.67 to 1.13; 1 RCT and 1 NRSI, 135 women; I^2^= 0%; low-certainty evidence) or fewer women will require transfusion (RR 0.82, 95% CI 0.74 to 0.91; 1 RCT and 4 NRSI; I^2^= 0%; very low-certainty evidence).

#### 1.3 Intraoperative visceral organ injury

Few cases of bladder injury were reported, therefore, we found similar event rates for both groups (RR 0.36, 95% CI 0.12 to 1.07; 1 RCT and 4 NRSI, 649 women; I^2^= 0%; low-certainty evidence). Potentially, the risk of bladder injury could be lower with conservative surgery (RR 0.32, 95% CI 0.19 to 0.54; 1 RCT and 6 NRSI; I^2^= 0%; very low-certainty evidence)(Figure 4). Due to the low event rate, the non-inferiority analysis was uninformative (Figures S4 and S5).

**Figure 4.** Comparison of uterine conservative surgery to hysterectomy on bladder injury in women with placenta accreta spectrum disorders. RCT, randomized controlled trial; IV, inverse variance; NRSI, nonrandomized study of intervention.

In the two studies that investigated ureter injuries, no cases were observed in either arm (2 NRSI, 87 women; very low-certainty evidence). Likewise, no cases of bowel injury were reported with conservative surgery, and seven were reported with hysterectomy (RR 0.17, 95% CI 0.02 to 1.65; 3 NRSI; I^2^= 17%; very low-certainty evidence).

#### 1.4 Reoperation

Very few cases of reoperation were reported in both groups, hence, the pooled estimate was very wide (RR 0.50, 95% CI 0.07 to 3.51; 1 RCT and 3 NRSI; I^2^= 0%; very low-certainty evidence)(Figure S6). The non-inferiority analysis was uninformative due to low event rates (Figures S7 and S8).

#### 1.5 Long-term complications of surgery

Vesicovaginal fistula formation was the only long-term complication reported across studies. As previously observed, this outcome was rare in both groups, leading to a wide effect estimate (RR 1.40, 95% CI 0.16 to 12.08; 1 RCT and 1 NRSI, 294 women; I² = 0%; very low-certainty evidence)(Figure S9). The non-inferiority analysis was uninformative due to low event rates (Figures S10 and S11).

#### 1.6 Duration of surgery

There was little to no difference in the duration of surgery (MD −36.31 minutes, 95% CI −73.06 to 0.43; 1 RCT and 2 NRSI, 147 women; I^2^= 54%; low-certainty evidence). Very uncertainly, conservative surgery could be shorter, however, high heterogeneity limits the confidence in this possibility (MD −26.45 minutes, 95% CI −45.35 to −7.56; 1 RCT and 6 NRSI; I^2^= 89%; very low-certainty evidence)(Figure 5).

**Figure 5.** Comparison of uterine conservative surgery to hysterectomy on the duration of surgery in women with placenta accreta spectrum disorders. RCT, randomized controlled trial; IV, inverse variance; NRSI, nonrandomized study of intervention.

#### 1.7 Duration of hospital stay

There was little to no difference in the duration of hospital stay (MD −0.39 days, 95% CI −0.87 to 0.08; 1 RCT and 4 NRSI; I^2^=33%; very low-certainty evidence). This could depend on institutional protocols and may not relate to the surgeries investigated.

#### 1.7 Admission to the intensive care unit (ICU)

There was little to no difference in the risk of ICU admission (RR 1.11, 95% CI 0.83 to 1.47; 1 RCT and 4 NRSI; I^2^= 0%; very low-certainty evidence).

### Certainty of evidence

The certainty of the evidence ranged from low to very low, according to the GRADE assessment (Table 2). Whenever the analysis included nonrandomized studies, the evidence was limited to low. To overcome these limitations on evidence, the Review Information Size (RIS) necessary to not downgrade for imprecision, for equally divided randomized controlled trials, would be: 166 to 594 participants for pRBC, and 210 to 4908 participants for blood loss, assuming moderate to small effects for the outcomes.

Conversely, more rare outcomes (visceral organ injury and ICU admission) would need far greater sample sizes, which might not be feasible.

## DISCUSSION

### Summary of main results

Conservative surgery has the advantage of preserving the uterus with surgical outcomes that were similar to hysterectomy in our analyses, in light of low-certainty evidence. Visceral organ injury, reoperation, and long-term complications were fairly rare, while blood loss was similar. In this regard, there is a high probability that conservative surgery is non-inferior to hysterectomy. However, patients had to have a specific PAS topography to be eligible for conservative surgery, with generally at least 2 cm of healthy myometrium on the anterior uterine wall, >50% of the uterine circumference free of PAS, without vesicouterine fibrosis, or extensive invasion of the lower anterior uterine wall.

### Limitations of the evidence included in the review

The most important limitation of our study is the inclusion of nonrandomized studies of interventions. Major limitations that future trials on PAS would face are the low incidence of PAS, which requires longer recruitment and results in smaller samples, the long-term follow-up needed to determine patient-centered outcomes, and the specific placenta topography that limits the number of eligible women to be recruited. Fundamentally, nonrandomized studies are better at evaluating potential harm than potential benefit^27^.

Inherent consequences of results drawn from these studies are uncontrolled confounding and suboptimal outcome reporting. Even after excluding high-risk studies from the main analysis, the confidence in the estimates provided was low. If only the data of the randomized trial were analyzed, the small sample size and wide confidence intervals would make it unreasonable to assume any given effect for conservative surgery.

Additionally, not all patients can be considered for conservative surgery, as there are topographical constraints to this technique. Another limitation was the lack of patient-driven outcomes in the included studies. However, it could be argued that uterus preservation (with reasonable safety) would be the most prominent outcome for patients.

### Agreements and disagreements with other studies or reviews

The success rate of conservative surgery in preserving fertility was reviewed elsewhere, and hence, was not one of our outcomes of interest^28^. Notably, subsequent pregnancy is possible, however, adverse maternal outcomes or recurrence of PAS was reported in nearly 20% of patients, and hysterectomy was necessary for around 2% of all women^28,29^. Such findings should be interpreted in the context of very low-certainty evidence due to the limitations of these studies.

Our results add substantially to the initial assessment from FIGO in 2018, in which it was suggested that leaving the placenta in situ was a valid management option for women desiring to preserve fertility while conservative surgery remained unclear^2^. Here we present both randomized and prospective nonrandomized data that suggest the safety of conservative surgery in some outcomes. In theory, considering both nonsurgical and surgical conservative management for PAS would be ideal for women desiring to preserve fertility, as these approaches have different shortcomings and eligibility criteria. Moreover, there are many variations of conservative surgery, which include prophylactic balloon occlusion or major vascular intervention (such as the triple P procedure)^30^. However, many of these studies did not compare their interventions to hysterectomy and, therefore, were excluded from our review. The studies that were eventually included were not randomized, and almost all had a high risk of bias, making it impossible to derive any conclusions about prophylactic vascular interventions. In previous investigations, meta-analyses and randomized controlled trials reported conflicting results for prophylactic balloon occlusion in women undergoing hysterectomy for PAS^31,32^.

In terms of intraoperative safety, the management of PAS requires careful planning due to the complexity of the procedures and the high risk of visceral organ injuries. Urological complications occur in approximately 20% of hysterectomies performed for PAS^33^, and massive blood loss remains both common and difficult to predict^34^. In our analysis, injury and transfusion rates appeared non-inferior between groups; however, strategies to effectively mitigate these intraoperative challenges remain elusive.

Moreover, in the non-inferiority analysis, we found a high probability that conservative surgery is non-inferior to hysterectomy in terms of blood loss. However, we could not make the same claim for other critical outcomes, such as intraoperative visceral organ injury and long-term complications. This limitation is largely attributable to the low prevalence of these events, which resulted in underpowered analyses. Nonetheless, it is unlikely that a potential increase in harm associated with conservative surgery would be detected.

### Implications for practice

Conservative surgery in women diagnosed with placenta accreta spectrum disorders, and with sufficiently healthy lower anterior uterine wall and uterine circumference, may not lead to higher blood loss, number of packs of red blood cells transfused, intraoperative visceral organ injury, and duration of surgery compared to hysterectomy. Therefore, uterus preservation is possible without the risk of substantial harm in critical outcomes likely increasing. This surgical procedure is not possible in all women diagnosed with PAS, especially in the presence of vesicouterine fibrosis.

### Implications for research

Further research should seek to increase the total sample size of randomized trials comparing conservative surgery to hysterectomy in order to minimize imprecision on confidence interval boundaries and review information size. Additionally, it would be ideal to determine whether there are specific variations of conservative surgery, especially involving prophylactic balloon occlusion, that result in better surgical outcomes. At present, our results suggest that conservative management does not result in substantially increased harm to women compared to hysterectomy, but more precise estimates would allow the determination of possible benefits.

## Data Availability

All data produced in the present study are available upon reasonable request to the authors

## DECLARATIONS

## Ethics approval and consent to participate

Not applicable

## Consent for publication

Not applicable

## Availability of data and materials

The data can be shared upon request

## Acknowledgments

Not applicable.

