## Supplementary material for "Conservative surgery vs hysterectomy for placenta accreta spectrum disorders: a systematic review and meta-analysis": Table S1 Characteristics of included studies (from another version of this preprint: version 2, 2025-04-25)

Nieto-Calvache 2024

| **Study characteristics** | |
|---|---|
| Methods | Multicenter study, parallel-group design<br>**Randomization**: Computer generated<br>**Blinding**: No<br>**Drop-outs**: Crossover of 9 participants in intervention group and 7 in hysterectomy group<br>**Number of participants**: 60<br>**Power analysis for sample size**: No<br>**Follow-up:** 6 weeks postpartum |
| Participants | **Eligibility for conservative surgery**: the bladder can be separated from the uterus, at least 2 cm of healthy myometrium caudal to the PAS area on the anterior uterine wall, and >50% of the uterine circumference free of PAS<br>**Exclusion criteria**: aged < 18 years, active bleeding, no placenta previa, and no previous live children |
| Intervention | **Intervention:** Conservative surgery (specific variation: One step surgery)<br>**Control group**: hysterectomy<br>**Preventive vascular intervention**: No |
| Outcomes | 1. Intraoperative bleeding volume<br>2. Length of surgery<br>3. Length of hospitalization<br>4. RBCU transfusion<br>5. Number of RBCU transfused<br>6. Bladder injury<br>7. Surgical reintervention<br>8. Admission to intensive care unit<br>9. Adverse event |

Nieto-Calvache 2023

| **Study characteristics** |
|---|

| | |
|---|---|
| Methods | Multicenter study, parallel-group, prospective non-randomized design<br><br>**Randomization**: No<br><br>**Blinding**: No<br><br>**Drop-outs**: Not reported<br><br>**Number of participants**: 75<br><br>**Power analysis for sample size**: No |
| Participants | **Eligibility for conservative surgery**: the bladder can be separated from the uterus, at least 2 cm of healthy myometrium caudal to the PAS area on the anterior uterine wall, and >50% of the uterine circumference free of PAS<br><br>**Exclusion criteria**: Patients with severe vesicouterine fibrosis (type 4 PAS on the topographic classification) or lower lateral placental involvement (low parametrial invasion or type 2 lower topographic classification) were excluded as, in those cases, it is not possible to perform OSCS |
| Intervention | **Intervention:** Conservative surgery (specific variation: One step surgery)<br><br>**Control group**: hysterectomy<br><br>**Preventive vascular intervention**: No |
| Outcomes | 1. Intraoperative bleeding volume<br><br>2. Length of surgery<br><br>3. Length of hospitalization<br><br>4. RBCU transfusion<br><br>5. Number of RBCU transfused<br><br>6. Bladder injury<br><br>7. Surgical reintervention |

Paping 2024

| **Study characteristics** | |
|---|---|
| Methods | Multicenter, parallel-group, prospectively collected-data, retrospective analysis, non-randomized design<br><br>**Randomization**: Computer generated<br><br>**Blinding**: No<br><br>**Drop-outs**: In the intention-to-treat analysis, 26 (46.4%) of prospectively assigned conservative surgery cases were instead |

| | |
|---|---|
| | managed by hysterectomy. Conversely, only 5 cases (3%) of hysterectomy were managed by conservative management. |
| | **Number of participants**: 224 |
| | **Power analysis for sample size**: No |
| Participants | **Eligibility for conservative surgery**: Based on each center's protocol. |
| | **Exclusion criteria**: - |
| Intervention | **Intervention:** Conservative surgery |
| | **Control group**: hysterectomy (third arm was leaving the placenta-in-situ, not included in the review) |
| | **Preventive vascular intervention**: No |
| Outcomes | 1. Intraoperative bleeding volume |
| | 2. Length of surgery |
| | 3. Length of hospitalization |
| | 4. RBCU transfusion |
| | 5. Number of RBCU transfused |
| | 6. Bladder injury |
| | 7. Surgical reintervention |
| | 8. Admission to intensive care unit |
| | 9. Adverse event |

Abouda 2024

| **Study characteristics** | |
|---|---|
| Methods | Single-center study, parallel-group, prospective, non-randomized design |
| | **Randomization**: No |
| | **Blinding**: No |
| | **Drop-outs**: 0 |
| | **Number of participants**: 12 |
| | **Power analysis for sample size**: No |
| Participants | **Eligibility for conservative surgery**: at least 2 cm of healthy myometrium caudal to the PAS area on the anterior uterine wall, and >50% of the uterine circumference free of PAS |

| | |
|---|---|
| | **Exclusion criteria**: successful manual removal of the placenta without significant bleeding and the absence of histologic confirmation of PAS postoperatively |
| Intervention | **Intervention:** Conservative surgery (specific variation: One-step surgery)<br>**Control group**: hysterectomy<br>**Preventive vascular intervention**: No |
| Outcomes | 1. Intraoperative bleeding volume (Hb change)<br>2. Number of pRBC transfused<br>3. Duration of surgery |

van Beekhuizen 2021

| Study characteristics | |
|---|---|
| Methods | Multicenter study, parallel-group, prospectively-collected data, retrospective analysis, non-randomized design<br>**Randomization**: No<br>**Blinding**: No<br>**Drop-outs**: Not informed<br>**Number of participants**: 278<br>**Power analysis for sample size**: No |
| Participants | **Eligibility for conservative surgery**: Not informed<br>**Exclusion criteria**: Not informed |
| Intervention | **Intervention:** Conservative surgery<br>**Control group**: hysterectomy (leaving the placenta in situ was the third arm, and extirpative management was the fourth arm, both not included in the review)<br>**Preventive vascular intervention**: No |
| Outcomes | 1. Intraoperative bleeding volume<br>2. Number of RBCU transfused<br>3. ICU admission<br>4. Intraoperative visceral organ injury<br>5. Reoperation |
| Observations | |

Vuong 2024

| **Study characteristics** | |
|---|---|
| Methods | Single-center study, parallel-group, retrospective, non-randomized design<br>**Randomization**: No<br>**Blinding**: No<br>**Drop-outs**: Retrospective design<br>**Number of participants**: 296<br>**Power analysis for sample size**: No |
| Participants | **Eligibility for conservative surgery**: pregnant women with a diagnosis of PAS at more than 28 weeks of pregnancy and more than 2 cm of healthy myometrium<br>**Exclusion criteria**: emergency management such as massive bleeding immediately before surgery, and uterine rupture. If a hysterectomy was indicated after the steps of the MOSCUS procedure proved insufficient, the patient was also excluded from the study. Other exclusion criteria included missing necessary data on the patient's file and the patient not receiving pathology results. |
| Intervention | **Intervention:** Resection surgery + preventive bilateral uterine artery ligation + B-lynch suture<br>**Control group**: hysterectomy<br>**Preventive vascular intervention**: No |
| Outcomes | 1. Intraoperative bleeding volume<br>2. Length of surgery<br>3. Length of hospitalization<br>4. RBCU transfusion<br>5. Number of RBCU transfused<br>6. Bladder injury<br>7. Surgical reintervention<br>8. Admission to intensive care unit |
| Observations | 13 hysterectomies and 25 conservative surgeries were considered emergency surgeries |

Aryananda 2023

| Study characteristics | |
|---|---|
| Methods | Single-center study, parallel-group, retrospective, non-randomized design<br>**Randomization**: No<br>**Blinding**: No<br>**Drop-outs**: Retrospective design<br>**Number of participants**: 126<br>**Power analysis for sample size**: No |
| Participants | **Eligibility for conservative surgery**: PAS topography above the bladder trigone (supero-anterior or upper parametrium PAS) was deemed suitable for OSCS. All women with a previous cesarean section and a placenta previa in the index pregnancy were identified as having risk factors for PAS and were screened by ultrasound.<br>**Exclusion criteria**: dehiscence of the uterine scar without abnormally adherent placenta, no signs of increased vascularity on the uteroplacental surface and uterine serosa during the surgery, and women in an unstable hemodynamic condition before surgery without adequate pre-surgical ultrasound. Emergency PAS cases with vaginal bleeding or uterine rupture but in a stable hemodynamic condition were not excluded if ultrasound examinations could be performed. |
| Intervention | **Intervention:** Conservative surgery (variation: one-step surgery)<br>**Control group**: hysterectomy<br>**Preventive vascular intervention**: No |
| Outcomes | 1. Intraoperative bleeding volume<br>2. RBCU transfusion<br>3. Number of RBCU transfused |
| Observations | 6 hysterectomies and 14 conservative surgeries were considered emergency surgeries |

Aryananda 2022

| Study characteristics | |
|---|---|
| Methods | Single-center study, parallel-group, retrospective, non-randomized design<br>**Randomization**: No<br>**Blinding**: No |

| | |
|---|---|
| | **Drop-outs**: Retrospective design<br>**Number of participants**: 234<br>**Power analysis for sample size**: No |
| Participants | **Eligibility for conservative surgery**: PAS topography above the bladder trigone (supero-anterior or upper parametrium PAS) was deemed suitable for OSCS. All women with a previous cesarean section and a placenta previa in the index pregnancy were identified as having risk factors for PAS and were screened by ultrasound.<br>**Exclusion criteria**: dehiscence of the uterine scar without abnormally adherent placenta, no signs of increased vascularity on the uteroplacental surface and uterine serosa during the surgery, and women in an unstable hemodynamic condition before surgery without adequate pre-surgical ultrasound. Emergency PAS cases with vaginal bleeding or uterine rupture but in a stable hemodynamic condition were not excluded if ultrasound examinations could be performed. |
| Intervention | **Intervention:** Conservative surgery (variation: MOSCUS)<br>**Control group**: hysterectomy<br>**Subgroup**: Resection surgery + bilateral iliac artery ligation<br>**Preventive vascular intervention**: No. Only a subgroup of patients underwent bilateral internal iliac artery ligation |
| Outcomes | 1. Intraoperative bleeding volume<br>2. Length of surgery<br>3. Length of hospitalization<br>4. RBCU transfusion<br>5. Number of RBCU transfused<br>6. Bladder injury<br>7. Surgical reintervention<br>8. Admission to intensive care unit |

Jafari 2022

| **Study characteristics** | |
|---|---|
| Methods | Single-center study, parallel-group, retrospective, non-randomized design<br>**Randomization**: No |

| | |
|---|---|
| | **Blinding**: No |
| | **Drop-outs**: Retrospective design |
| | **Number of participants**: 68 |
| | **Power analysis for sample size**: No |
| Participants | **Eligibility for conservative surgery**: Not informed |
| | **Exclusion criteria**: Not informed |
| Intervention | **Intervention:** Conservative surgery |
| | **Control group**: hysterectomy |
| | **Preventive vascular intervention**: No |
| Outcomes | 1. Hospital stay |
| | 2. RBCU transfusion |
| | 3. Duration of surgery |
| Observations | |

Durukan 2021

| **Study characteristics** | |
|---|---|
| Methods | Single-center study, parallel-group, retrospective, non-randomized design |
| | **Randomization**: No |
| | **Blinding**: No |
| | **Drop-outs**: Retrospective design |
| | **Number of participants**: 65 |
| | **Power analysis for sample size**: No |
| Participants | **Eligibility for conservative surgery**: Not informed |
| | **Exclusion criteria**: Not informed |
| Intervention | **Intervention:** Conservative surgery |
| | **Control group**: hysterectomy |
| | **Preventive vascular intervention**: No |
| Outcomes | 1. Intraoperative bleeding volume |
| | 2. RBCU transfusion |
| | 3. ICU stay |
| Observations | |

Üstünyurt 2020

| **Study characteristics** | |
|---|---|
| Methods | Single-center study, parallel-group, retrospective, non-randomized design<br>**Randomization**: No<br>**Blinding**: No<br>**Drop-outs**: Retrospective design<br>**Number of participants**: 97<br>**Power analysis for sample size**: No |
| Participants | **Eligibility for conservative surgery**: sonographic and intraoperative findings are appropriate in patients who prefer the preservation of the uterus, uterine sparing surgery is performed<br>**Exclusion criteria**: Conservative surgery is not performed in cases with invasion into the parametrium and/ or cervix and invasion of more than 50% of the anterior surface of the uterus |
| Intervention | **Intervention:** Conservative surgery<br>**Control group**: hysterectomy<br>**Preventive vascular intervention**: No |
| Outcomes | 1. Intraoperative bleeding volume<br>2. RBCU transfusion<br>3. Number of RBCU transfused<br>4. Duration of surgery<br>5. Duration of hospital stay<br>6. ICU admission<br>7. Intraoperative visceral organ injury |
| Observations | Statistical analysis was adjusted for confounders such as age, gravidity, parity, previous cesarean delivery, curettage, and myomectomy |

Kilicci 2018

| **Study characteristics** | |
|---|---|
| Methods | Single-center study, parallel-group, retrospective, non-randomized design<br>**Randomization**: No |

| | |
|---|---|
| | **Blinding**: No |
| | **Drop-outs**: Retrospective design |
| | **Number of participants**: 51 |
| | **Power analysis for sample size**: No |
| Participants | **Eligibility for conservative surgery**: (a) Presence of an uninvolved area close to uterine arteries, (b) Applicability of urinary bladder dissection from anterior uterine wall |
| | **Exclusion criteria**: Patients undergoing urgent Cesarean section |
| Intervention | **Intervention:** Conservative surgery |
| | **Control group**: hysterectomy |
| | **Preventive vascular intervention**: No |
| Outcomes | 1. Number of RBCU transfused<br>2. Duration of surgery<br>3. Duration of hospital stay |
| Observations | |
