## Supplementary material for "Conservative surgery vs hysterectomy for placenta accreta spectrum disorders: a systematic review and meta-analysis": Table S2 Risk of bias of included studies (from another version of this preprint: version 2, 2025-04-25)

Nieto-Calvache 2024

| **Risk of bias (Rob 2)** | | |
|---|---|---|
| Bias | Author's judgement | Supporting information |
| Random sequence generation (selection bias) | Low risk | Computer generated |
| Allocation concealment (selection bias) | Low risk | An appropriate analysis was used to estimate the effect of assignment to intervention |
| Incomplete outcome data (attrition bias) | Low risk | Outcome data were available for all, or nearly all, randomized participants |
| Measurement of the outcome | Some concerns | Open-label study. It is unlikely that assessment of the surgical outcomes was influenced by knowledge of intervention received. |
| Selective reporting (re-porting bias) | Low risk | Study conducted according to a prospectively registered protocol |

Nieto-Calvache 2023

| **Risk of bias (ROBINS-I)** | | |
|---|---|---|
| Bias | Author's judgement | Supporting information |
| Bias due to confounding | Some concerns | Comparable baseline characteristics, BMI and number of previous cesareans not reported. Adjusted analysis not performed. |
| Bias in the selection of participants into the study | Some concerns | Prospectively recruited study, however, intervention could change based on intraoperative assessment. |
| Bias in classification of the intervention | Low risk | Well defined intervention status. |
| Bias due to deviations from intended interventions | Low risk | There weren't substantial deviations. |
| Bias due to missing outcome data | Some concerns | Complete case analysis. |

| | | |
|---|---|---|
| Bias in measurement of the outcome | Some concerns | The methods of outcome assessment were comparable across intervention groups and outcome measure is only minimally affected by knowledge of the intervention |
| Bias in selection of the reported result | Some concerns | There was no protocol. However, there appears to be no issues with multiple analyses or subgroups and nearly all relevant surgical outcomes were reported |

Paping 2024

| **Risk of bias (ROBINS-I)** | | |
|---|---|---|
| Bias | Author's judgement | Supporting information |
| Bias due to confounding | Some concerns | Comparable baseline characteristics, BMI not reported. Adjusted analysis not performed. |
| Bias in the selection of participants into the study | Low risk | Prospectively recruited study. "Participants were first grouped based on the actual treatment performed after intraoperative assessment and staging. A second analysis was performed using an intention-to-treat analysis, whereby participants were grouped based on the preoperative planned surgery". |
| Bias in classification of the intervention | Low risk | Well defined intervention status. |
| Bias due to deviations from intended interventions | Low risk | There weren't substantial deviations. |
| Bias due to missing outcome data | Low risk | Nearly all patients had outcome data. Excluded cases were not related to lack of data. |
| Bias in measurement of the outcome | Some concerns | The methods of outcome assessment were comparable across intervention groups and outcome measure is only minimally affected by knowledge of the intervention |
| Bias in selection of the reported result | Some concerns | There was no protocol. However, there appears to be no issues with multiple analyses or subgroups and nearly all relevant surgical outcomes were reported |

Abouda 2024

| **Risk of bias (ROBINS-I)** |
|---|

| Bias | Author's judgement | Supporting information |
|---|---|---|
| Bias due to confounding | Some concerns | Comparable baseline characteristics. Adjusted analysis not performed. |
| Bias in the selection of participants into the study | Low risk | Prospectively recruited study according to a prespecified protocol. |
| Bias in classification of the intervention | Low risk | Well defined intervention status. |
| Bias due to deviations from intended interventions | Low risk | There weren't substantial deviations. |
| Bias due to missing outcome data | Low risk | Outcome data was complete. |
| Bias in measurement of the outcome | Low risk | The methods of outcome assessment were comparable across intervention groups and outcome measure is only minimally affected by knowledge of the intervention. A prespecified protocol defined how outcome would be measured before the start of the study. |
| Bias in selection of the reported result | Some concerns | Study conducted according to protocol. |

van Beekhuizen 2021

| **Risk of bias (ROBINS-I)** | | |
|---|---|---|
| Bias | Author's judgement | Supporting information |
| Bias due to confounding | Some concerns | Comparable baseline characteristics, BMI and maternal age not reported. Adjusted analysis not performed. |
| Bias in the selection of participants into the study | Some concerns | Prospectively recruited study, however, intervention could change based on intraoperative assessment. |
| Bias in classification of the intervention | Low risk | Well defined intervention status. |
| Bias due to deviations from intended interventions | Low risk | There weren't substantial deviations. |

| | | |
|---|---|---|
| Bias due to missing outcome data | Low risk | Nearly all patients had outcome data. Excluded cases were not related to lack of data. |
| Bias in measurement of the outcome | Some concerns | The methods of outcome assessment were comparable across intervention groups and outcome measure is only minimally affected by knowledge of the intervention |
| Bias in selection of the reported result | Some concerns | There was no protocol. However, there appears to be no issues with multiple analyses or subgroups and nearly all relevant surgical outcomes were reported |

Vuong 2024

| **Risk of bias (ROBINS-I)** | | |
|---|---|---|
| Bias | Author's judgement | Supporting information |
| Bias due to confounding | High | Retrospective analysis, not adjusted. |
| Bias in the selection of participants into the study | High | Retrospective recruited study, intervention could change based on intraoperative assessment. |
| Bias in classification of the intervention | Low risk | Well defined intervention status. |
| Bias due to deviations from intended interventions | Low risk | There weren't substantial deviations. |
| Bias due to missing outcome data | Some concerns | Complete case analysis. |
| Bias in measurement of the outcome | Some concerns | The methods of outcome assessment were comparable across intervention groups and outcome measure is only minimally affected by knowledge of the intervention |
| Bias in selection of the reported result | Some concerns | There was no protocol. However, there appears to be no issues with multiple analyses or subgroups and nearly all relevant surgical outcomes were reported |

Aryananda 2023

| **Risk of bias (ROBINS-I)** | | |
|---|---|---|
| Bias | Author's judgement | Supporting information |

| | | |
|---|---|---|
| Bias due to confounding | High | Retrospective analysis, not adjusted. |
| Bias in the selection of participants into the study | High | Retrospective recruited study, intervention could change based on intraoperative assessment. |
| Bias in classification of the intervention | Some concerns | A substantial number of failed conservative management may have crossed over to hysterectomy, compromising the analysis |
| Bias due to deviations from intended interventions | Low | There weren't substantial deviations. |
| Bias due to missing outcome data | Some concerns | Complete case analysis. |
| Bias in measurement of the outcome | Some concerns | The methods of outcome assessment were comparable across intervention groups and outcome measure is only minimally affected by knowledge of the intervention. |
| Bias in selection of the reported result | High | There was no protocol. Important outcomes were not reported. |

Aryananda 2022

| **Risk of bias (ROBINS-I)** | | |
|---|---|---|
| Bias | Author's judgement | Supporting information |
| Bias due to confounding | High | Retrospective analysis, not adjusted. |
| Bias in the selection of participants into the study | Some concerns | Retrospective recruited study, but consecutive cases were included. A flow diagram of patient selection is provided. Selection criteria was reasonable to assume high risk was not present (only cases of upper trigonal bladder invasion were included in hysterectomy and conservative surgery). |
| Bias in classification of the intervention | Low risk | Well defined intervention status. |
| Bias due to deviations from intended interventions | Low risk | There weren't substantial deviations. |

| | | |
|---|---|---|
| Bias due to missing outcome data | Some concerns | Complete case analysis. |
| Bias in measurement of the outcome | Some concerns | The methods of outcome assessment were comparable across intervention groups and outcome measure is only minimally affected by knowledge of the intervention |
| Bias in selection of the reported result | Some concerns | There was no protocol. However, there appears to be no issues with multiple analyses or subgroups and nearly all relevant surgical outcomes were reported |

Jafari 2022

| **Risk of bias (ROBINS-I)** | | |
|---|---|---|
| Bias | Author's judgement | Supporting information |
| Bias due to confounding | High | Retrospective analysis, not adjusted. |
| Bias in the selection of participants into the study | High | Retrospective recruited study, intervention could change based on intraoperative assessment. |
| Bias in classification of the intervention | Low | Classification was well defined. |
| Bias due to deviations from intended interventions | Low | There weren't substantial deviations. |
| Bias due to missing outcome data | Some concerns | Complete case analysis. |
| Bias in measurement of the outcome | Some concerns | The methods of outcome assessment were comparable across intervention groups and outcome measure is only minimally affected by knowledge of the intervention. |
| Bias in selection of the reported result | Some concerns | There was no protocol. Nearly all important outcomes were reported. |

Jafari 2022

| **Risk of bias (ROBINS-I)** | | |
|---|---|---|
| Bias | Author's judgement | Supporting information |

| | | |
|---|---|---|
| Bias due to confounding | High | Retrospective analysis, not adjusted. |
| Bias in the selection of participants into the study | High | Retrospective recruited study, intervention could change based on intraoperative assessment. |
| Bias in classification of the intervention | Low | Classification was well defined. |
| Bias due to deviations from intended interventions | Low | There weren't substantial deviations. |
| Bias due to missing outcome data | Some concerns | Complete case analysis. |
| Bias in measurement of the outcome | Some concerns | The methods of outcome assessment were comparable across intervention groups and outcome measure is only minimally affected by knowledge of the intervention. |
| Bias in selection of the reported result | High | There was no protocol. Important outcomes were not reported. |

Durukan 2021

**Risk of bias (ROBINS-I)**

| Bias | Author's judgement | Supporting information |
|---|---|---|
| Bias due to confounding | High | Retrospective analysis, not adjusted. |
| Bias in the selection of participants into the study | High | Retrospective recruited study, intervention could change based on intraoperative assessment. |
| Bias in classification of the intervention | Some concerns | Conservative surgery may have been attempted in hysterectomy cases. |
| Bias due to deviations from intended interventions | Low | There weren't substantial deviations. |
| Bias due to missing outcome data | High | No information is provided |
| Bias in measurement of the outcome | Some concerns | The methods of outcome assessment were comparable across intervention groups and outcome measure is only minimally affected by knowledge of the intervention. |

| | | |
|---|---|---|
| Bias in selection of the reported result | High | There was no protocol. Important outcomes were not reported. |

Üstünyurt 2020

| **Risk of bias (ROBINS-I)** | | |
|---|---|---|
| Bias | Author's judgement | Supporting information |
| Bias due to confounding | Some concerns | Retrospective analysis, adjusted. |
| Bias in the selection of participants into the study | High | Retrospective recruited study, intervention could change based on intraoperative assessment. |
| Bias in classification of the intervention | Low | Classification was well defined. |
| Bias due to deviations from intended interventions | Low | There weren't substantial deviations. |
| Bias due to missing outcome data | Low | Patients were not excluded based on missing outcome data |
| Bias in measurement of the outcome | Some concerns | The methods of outcome assessment were comparable across intervention groups and outcome measure is only minimally affected by knowledge of the intervention. |
| Bias in selection of the reported result | Low | Nearly all outcomes of interest reported. there appears to be no selection based on outcome, intervention, multiple analyses, or different subgroups. |

Kilicci 2018

| **Risk of bias (ROBINS-I)** | | |
|---|---|---|
| Bias | Author's judgement | Supporting information |
| Bias due to confounding | High | Retrospective analysis, not adjusted. |
| Bias in the selection of participants into the study | Some concerns | Retrospective recruited study, intervention could change based on intraoperative assessment. Selection criteria was adequate. |

| | | |
|---|---|---|
| Bias in classification of the intervention | Low | Classification was well defined. |
| Bias due to deviations from intended interventions | Low | There weren’t substantial deviations. |
| Bias due to missing outcome data | Some concerns | Complete case analysis. |
| Bias in measurement of the outcome | Some concerns | The methods of outcome assessment were comparable across intervention groups and outcome measure is only minimally affected by knowledge of the intervention. |
| Bias in selection of the reported result | Some concerns | Not all outcomes of interest reported. |
